# Time to Recovery and Predictors of Severe Pneumonia in Children Under Five Years of Age at Referral and General Hospitals in Sidama Region, Southern Ethiopia, multicenter prospective cohort study

**DOI:** 10.1101/2025.05.13.25327555

**Authors:** Fantahun Funte Ware, Nana Chea Hankalo, Ruth Abriham Tadesse

## Abstract

**Background:** A longer period of hospitalization is known to raise the risk of local and systemic infection, which can lead to complicated pneumonia. These negative impacts of inpatient management of severe pneumonia become more pronounced if the length of recovery time is prolonged. Therefore, the aim of this study was to assess the time to recovery from severe pneumonia and its predictors among under-five children.

**Methodology:** A multicenter prospective cohort study was conducted on 286 children under five years of age with severe pneumonia from March to July 2024 at referral and general hospitals in the Sidama Region, South Ethiopia. Data collectors received two-days training. Systematic sampling was employed to select participants. Data analysis was performed using STATA version 16, with Cox proportional regression. The Cox proportional hazard model assumption was validated using the Schoenfeld residual global test, which yielded a P-value of 0.505.

**Results:** Among 286 under-five children with severe pneumonia, the median time to recovery was 5 days (interquartile range = 4-7). The incidence of recovery rate was 17.27 (95% CI, 15.3- 19.4) per 100 person-days. Severe acute malnutrition (adjusted hazard ratio; 0.567, 95% CI (0.357- 0.90)), presence of danger signs (adjusted hazard ratio; 0.497, 95% CI (0.314-0.787)) and presence of co-morbidities (adjusted hazard ratio; 0.618, 95% CI (0.477- 0.800)) were associated with time to recovery from severe pneumonia in children younger than five years.

**Conclusions and recommendations:** The median recovery time of pediatric patients admitted with severe pneumonia was relatively high. Malnutrition, presence of danger signs, and comorbidities were significant associated factors; therefore, to reduce time to recovery from severe pneumonia by enhancing nutritional status, early detection and treatment of the danger signs, and comorbidity diseases.

## INTRODUCTION

Pneumonia, an acute respiratory infection affecting the lungs and caused by various pathogens including bacteria, viruses, and fungi, is a serious disease with high morbidity and mortality rates, particularly in pediatric populations, where it accounts for 65.2% of hospitalizations (1,2). Severe pneumonia, characterized by symptoms such as coughing, dyspnea, tachypnea, and general danger signs like an inability to drink, stridor in a calm child, or an oxygen saturation of less than 90% in room air, is a significant global health concern, it is responsible for 896,000 deaths out of 5.6 million in 2016 and 14% of all deaths in children under 5 years old, totaling 740,180 deaths in 2019, With an annual incidence of approximately 152 million cases, mostly in low- and middle-income countries, 8.7% of these cases progress to severe pneumonia requiring hospitalization (3–7).

Africa loses thousands of children due to pneumonia each year, which cause around 750,000 child deaths per year in sub-Saharan African countries (8). In Ethiopia, pneumonia is the leading causes of child mortality and morbidity and accounts for 18% of childhood deaths (9).

Long hospital duration of children experiences psychosocial problems of children and family, burdening hospital services in developing countries by increasing hospital bed occupancy capacity for inpatient (10,11). Treatment in the hospital for children with severe pneumonia places a significant burden on children and their families, including substantial expense and decreased quality of life (12). The main difficulty is the increasing duration of stay in the hospital and the poor response to antibiotic treatments, which has resulted in a decrease in pneumonia patient survival; A longer period of hospitalization is known to raise the risk of local and systemic infection, which can lead to complicated pneumonia; it also has huge economic disadvantages, either directly through medical expenses or indirectly through the loss of working hours suffered by parents of sick children; These negative impacts of inpatient management of severe pneumonia become more pronounced if the length of recovery time is prolonged (13,14).

Pneumonia continues to be a common cause of mortality and prolonged hospitalization of children in Ethiopia, which calls for innovative strategies that will come through systematic research to end the problems. Data on the time to recovery of severe pneumonia and its associated factors are important for planning child health care services but little is known about it, especially for under five children, and the title is not done by prospective multicenter cohort study design in study area as far as our search. Therefore, the aim is to assess time to recovery and its predictors of severe pneumonia among under-five children at Referral and General Hospital in Sidama Region.

## METHODS AND MATERIALS

### Study design, Study area and period

Multicenter prospective cohort was conducted in the Sidama National Regional State, which is located in the southern part of Ethiopia. The Central Statistical Agency (CSA) estimates the population of the region at 4,569,336. From these, children under 1 year constitute 3.19% of the population, children under 2 years comprise 5.18%, and children under 5 years 15.5%. The study settings were Referral and General Hospital (HUCSH, Adare General Hospital, Tula General Hospital, Leku General Hospital, Yirgalem General Hospital, Bona General Hospital, and Bensa Daye General Hospital) in the Sidama Region from March 22; 2024 to July 4; 2024. The follow up started on March 22, 2024 and ended on July 4, 2024.

### Source population

All children under five years of age diagnosed with pneumonia and admitted to pediatric ward of referral and General hospital in Sidama region, 2024

### Study population

All children under-five years of age diagnosed with severe pneumonia and admitted to inpatient pediatric ward of Referral and General hospital in Sidama region from March 22; 2024 to July 4; 2024

### Eligibility criteria

#### Inclusion criteria

All children aged from 1-59 months diagnosed with severe pneumonia and admitted to inpatient pediatrics ward of Referral and General Hospital in Sidama Region from March 22; 2024 to July 4; 2024

#### Exclusion criteria

Children less than one month or inpatient who admitted to NICU Children who have not guardians from March 22; 2024 to July 4; 2024

### Sample size determination

The sample size was determined using STATA software version 16 using power analysis for the Cox proportional hazard model by considering the following assumptions: 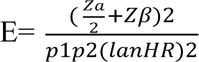 and 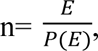 a variability standard deviation (SD) of 0.5, probability of withdrawal 0.1, HR is the hazard ratio of presence of danger signs was 1.46, and the probability of an event (recovery) was 0.94 from a previous study that was conducted in East Wallaga zone public hospitals, western Ethiopia (13). Finally, by adding 10% for lost to follow up, then the final **sample size was become 286**.

### Sampling technique and sampling procedure

The final sample size was allocated in each hospital using a proportional allocation formula based on the number of severe pneumonia. After allocation, the sampling interval of k value was calculated as the total number of children admitted with severe pneumonia at referral and general hospital divided by the total sample size (404/286 = ∼1.5) then k value becomes ∼2. Every second participant was selected after the first eligible participant using the lottery method.

### Variables

#### Dependent variables

Time to recovery of severe pneumonia

#### Independent variables

**Socio-demographic characteristics:** level of institutions, age, sex, residence, history of smoker in parents, housing condition and number of households

**Nutritional factors and immunization:** nutritional status, breast feeding status and Immunization status

**Pre-existing medical conditions and co-morbidities**: type of severe pneumonia, history of measles, HIV sero-status, history of URTI and other co-morbidities

**Factors related to duration of illness**: duration of illness before admission

### Data collection tools and technique

The data extraction format was developed from different previous studies(5,13,15–21). Data was collected by Seven BSC nurses by face to face interviews and reviewing the medical records using Sidamu afoo and Amharic language which translated from English version structured questionnaire and back to English to check the consistency. The questionnaire includes four parts: Part one include socio-demographic characteristics, Part two covers nutritional factors and immunization status, Part three includes preexisting medical or comorbidity condition, and part four includes duration of illness prior to admission.

### Data quality and management

The English version of the questionnaire is translated into Sidamu afoo and Amharic language for the better understanding by the respondents. A Pretest of the questionnaire was done on 5 % (14) respondents or study participants at Aleta Wondo General Hospital. Two days training was provided to data collectors. Supervision was done and the collected information was checked for its consistency & completeness by the principal investigators.

### Data management and analysis

The collected data were coded and entered into the Epi-data statistical software package version 4.6 and exported and analyzed by STATA version 16 for analysis. Descriptive statistics were calculated and summarized using text, tables, charts, and graphs. The median recovery times were determined and explained using the interquartile range (IQR).

Bivariate and multivariable Cox regression analyses were performed using the Cox proportional hazard regression model to control confounding variables to identify its predictors of time to recovery in children with severe pneumonia. Kaplan-Meier survival estimation was performed to determine the probability of survival. In this study proportional hazard assumption test was checked by using the Schoenfeld residual global test. The Schoenfeld residual global test P-value was 0.505, which met the Cox proportional hazard model assumption. Based on bivariate analysis, those variables having a p-value ***<*** 0.25 in the bivariate cox proportional hazard regression analysis were transferred to the multivariable cox proportional hazard regression analysis, and finally in multivariable cox proportional hazard regression analysis those variables with a P-value ***<*** 0.05 at 95 % confidence level were considered as an independent predictors of time to recovery of under five patients with severe pneumonia. Finally, presented by texts, tables, charts and graphs

### Operational definitions

**Treatment outcome:** Result was obtained after the use of a drug (22).

**Time to recovery:** the time from the date of admission to the event/ recovery clinically from severe pneumonia as declared by the physician during the study period (23).

**Recovery:** patients improved from severe pneumonia as declared by the physician (23)**. Event:** refers to recovery from an illness during the study period (24).

**Censored:** refers to children referred, died or discharged (unknown outcome) for any reason without recovery during the study period (24).

## RESULTS

### Socio demographic results

Among the 286 cases analyzed, around half, 141 (49.3%), were male, and 145 (59.1%) were female; 114 (40%) lived in urban areas, and 172 (60%) lived in rural areas, and the median age of cases analyzed was 12 months. 117 (41%) of children lived in a house that was not separated from the kitchen, and 25 (8.7%) had a family history of smoking.

**Table 1.** socio demographic characteristics of respondents, 2024.

| Variables |  | Frequency | Percent |
| --- | --- | --- | --- |
| Level of institution | Referral | 30 | 10.5 |
|  | General | 256 | 89.5 |
| Age ( in months) | 1 up to 12 | 151 | 52.8 |
|  | 13 up to 24 | 71 | 24.8 |
|  | 25 up to 36 | 34 | 11.9 |
|  | 37 up to 59 | 30 | 10.5 |
| Sex | Male | 141 | 49.3 |
|  | Female | 145 | 50.7 |
| Residence | Urban | 114 | 40 |
|  | Rural | 172 | 60 |
| Housing condition | Separated from kitchen | 169 | 59 |
|  | Not separated from kitchen | 117 | 41 |
| Family history of smoking | Yes | 25 | 8.7 |
|  | No | 261 | 91.3 |
| Family size | <5 | 180 | 63 |
|  | >= 5 | 106 | 37 |

### Nutritional; immunization and breast-feeding characteristics

Out of 286 analyzed, 120(42%) were well-nourished; 91(32%) were mild acute nutrition; 41(14%) were moderate acute malnutrition and 34(12%) were severe acute malnutrition.

**Table 2.** Nutritional, immunization and breast feeding characteristics, 2024.

| Variables |  | Frequency | Percent |
| --- | --- | --- | --- |
| **Nutritional status** | Well nourished | 120 | 42 |
|  | MAM | 91 | 32 |
|  | MOM | 41 | 14 |
|  | SAM | 34 | 12 |
| **Immunization status** | Fully immunized | 122 | 42.7 |
|  | Partially immunized | 50 | 17.5 |
|  | Immunized for age | 93 | 32.5 |
|  | Not immunized at all | 21 | 7.4 |
| **Breast feeding status** | Exclusive breast feeding | 194 | 67.8 |
|  | Not exclusive breast feeding | 92 | 32.2 |

### Pre-existing medical conditions and co morbidities characteristics at admission

Among 286 patients analyzed, 256(89.5%) had severe pneumonia and 30(10.5%) had very severe pneumonia; 147(51.4) had comorbidities.

**Table 3.** presence of danger signs; Preexisting medical conditions and co morbidities characteristics of under five years at referral and general Hospitals in Sidama Region Ethiopia, 2024.

| Variables |  | Frequency | Percent |
| --- | --- | --- | --- |
| Presence of danger signs | No | 256 | 89.5 |
|  | Yes | 30 | 10.5 |
| Comorbidities | Yes | 147 | 51.4 |
|  | No | 139 | 48.6 |
| Measles | Yes | 49 | 17.1 |
|  | No | 237 | 82.9 |
| Duration of illness before admission | 1 up to 3 days | 143 | 50 |
|  | 4 up to 7 days | 137 | 48 |
|  | 8 >_days | 6 | 2 |
| History of URTI | Yes | 92 | 32 |
|  | No | 194 | 68 |
| HIV sero Status | Yes | 1 | 0.3 |
|  | No | 285 | 99.7 |

### Treatment outcome of severe pneumonia

Among 286 cases analyzed of which 263(92%) were improved, 11(3.8%) were died and 12(4.2%) had unknown outcomes (self against medical services and referred).

**Figure 1.**
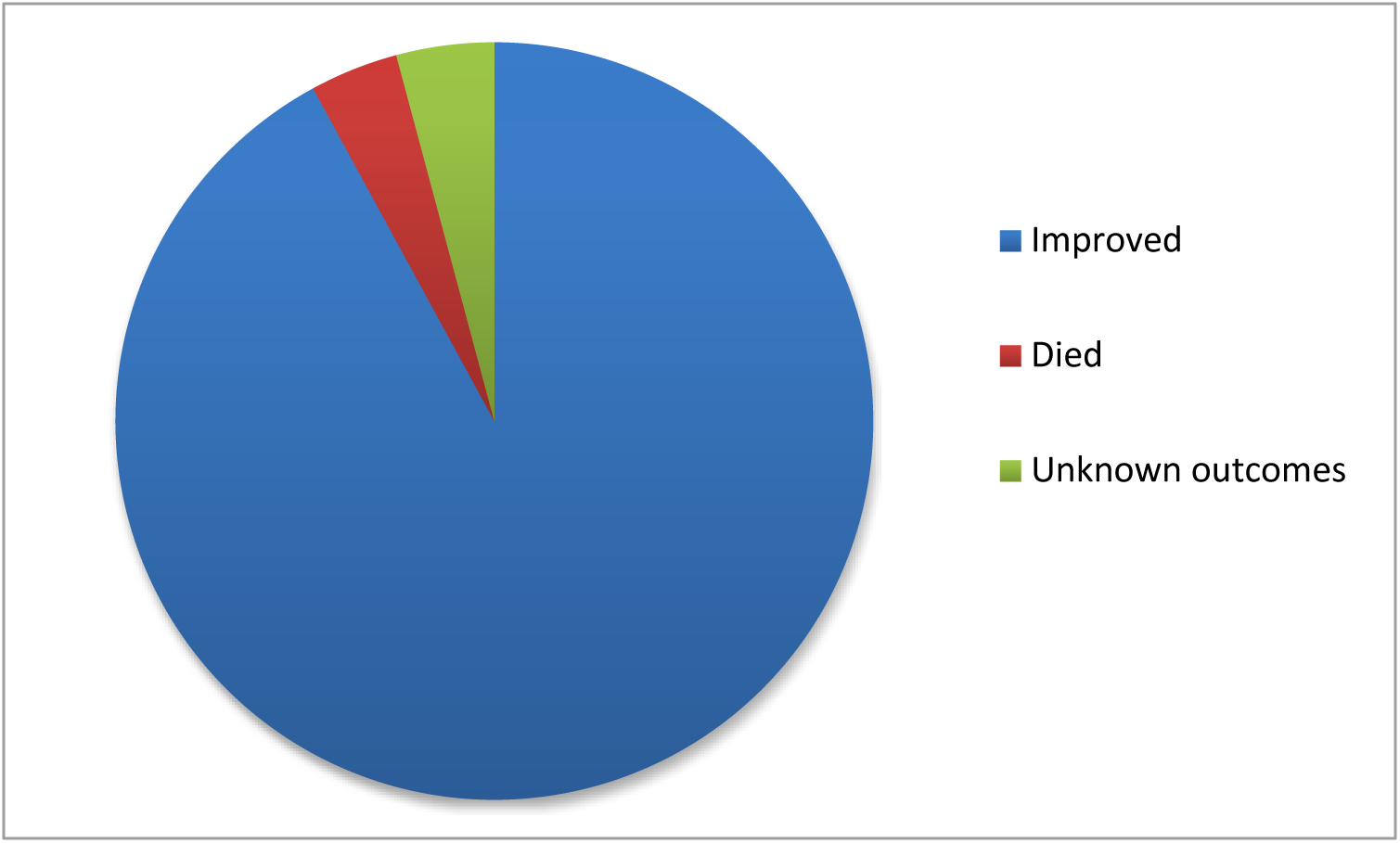
Treatment outcome of severe pneumonia of under five children in referral and general hospitals in Sidama Region (N=286).

### Kaplan–Meier Survival Curve

The median survival time to recovery from severe pneumonia among under-five children was found to be 5 days (IQR=4-7). The incidence of recovery rate was 17.27 (95% CI, 15.3, 19.4) per 100 person-days. The total person-days contributed by the study participants were 1523. The chance of survival was higher on the first day after a severe pneumonia diagnosis, but it decreased as follow-up time increased.

**Figure 2.**
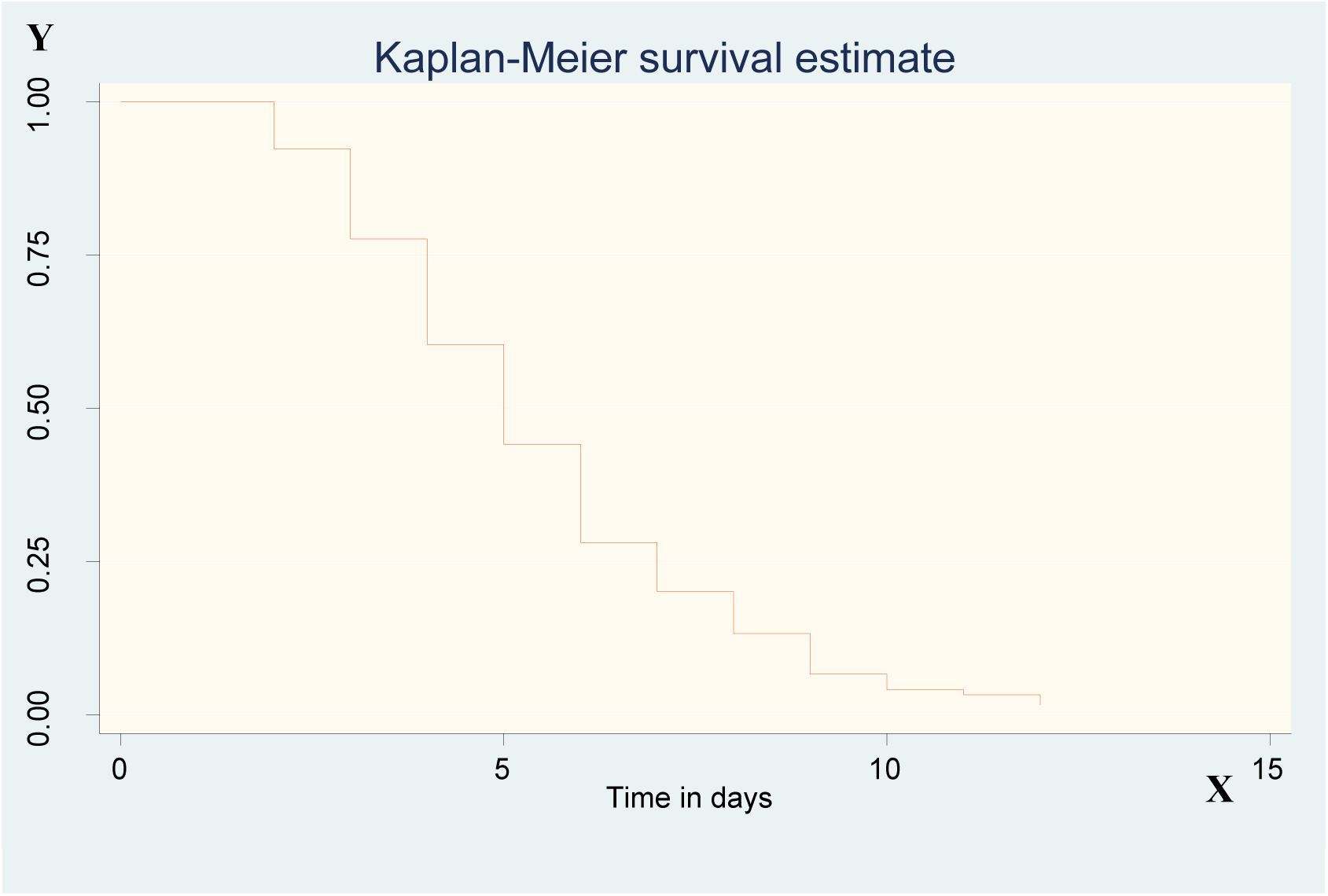
Overall Kaplan–Meier estimation of survival function from admission to recovery among under five severe pneumonia children admitted at the referral and general hospital in Sidama region, Ethiopia, 2024.

### Associated factors of time to recovery of severe pneumonia

Using the criteria of P value < 0.25 among children under five with severe pneumonia, the following variables were included in the multivariable Cox proportional hazard regression: nutritional status, immunization status, history of upper respiratory tract infection, breastfeeding status, presence of danger signs, housing condition, measles, and comorbidities.

Finally, nutritional status, presence of danger signs and comorbidities were statistically significant associated factors of time to recovery of severe pneumonia.

The hazard ratio of recovery of children who were admitted with severe acute malnutrition reduced by 43.3% compared to those admitted with well-nourished (AHR; 0.567, 95% CI (0.357, 0.90)). The hazard ratio of recovery of children who were admitted with moderate acute malnutrition reduced by 34.5% compared to those admitted with well-nourished (AHR; 0.655, 95% CI (0.443, 0.968)). The hazard ratio of recovery of children who were admitted with the presence of danger signs decreased by 50.3% compared to those admitted without danger signs (AHR; 0.497, 95% CI (0.314, 0.787)). The hazard ratio of recovery of children admitted with co- morbidity decreased by 38.2% as compared to those children who had no co-morbidity (AHR; 0.618, 95% CI (0.477, 0.800).

**Table 4.**
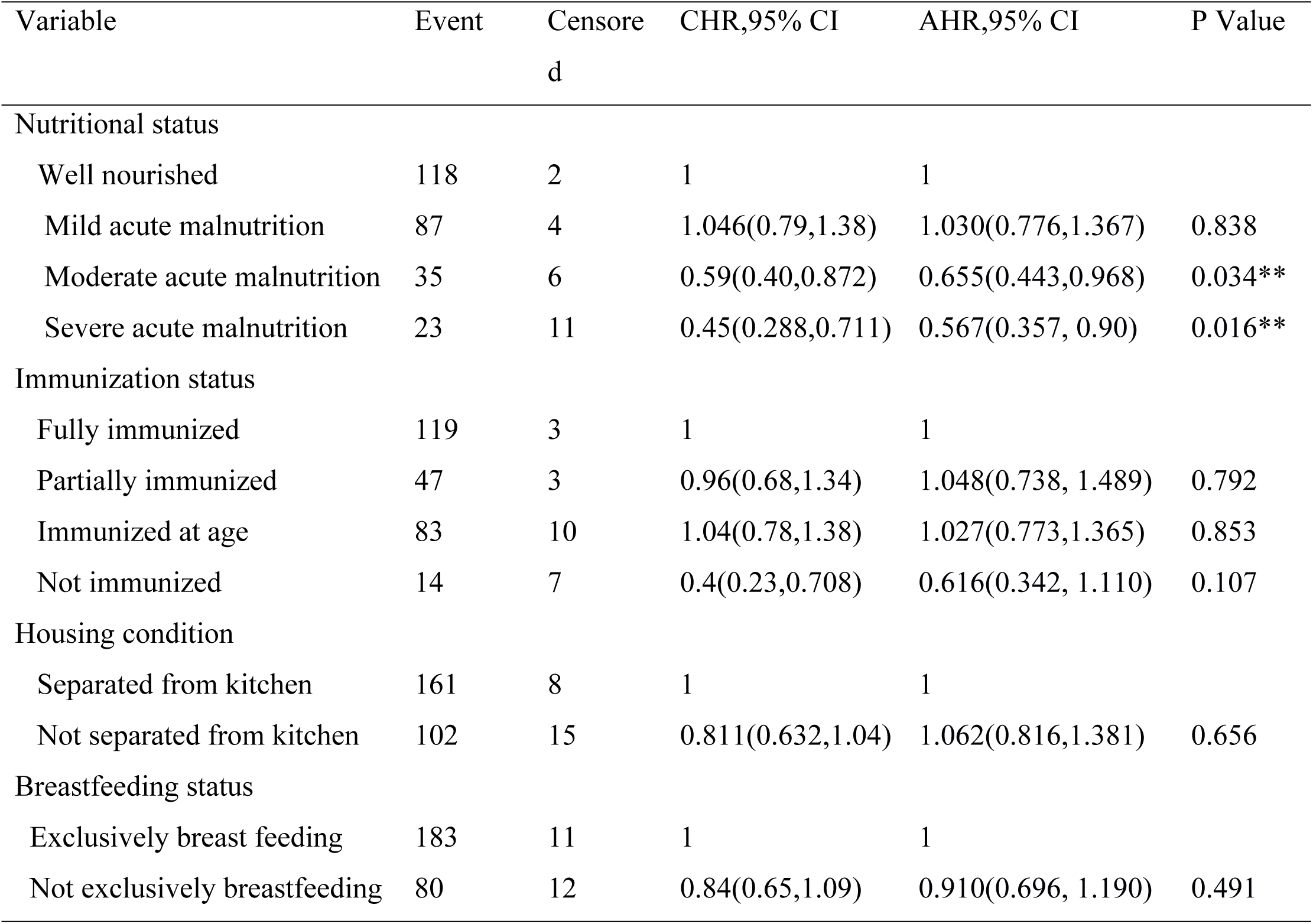
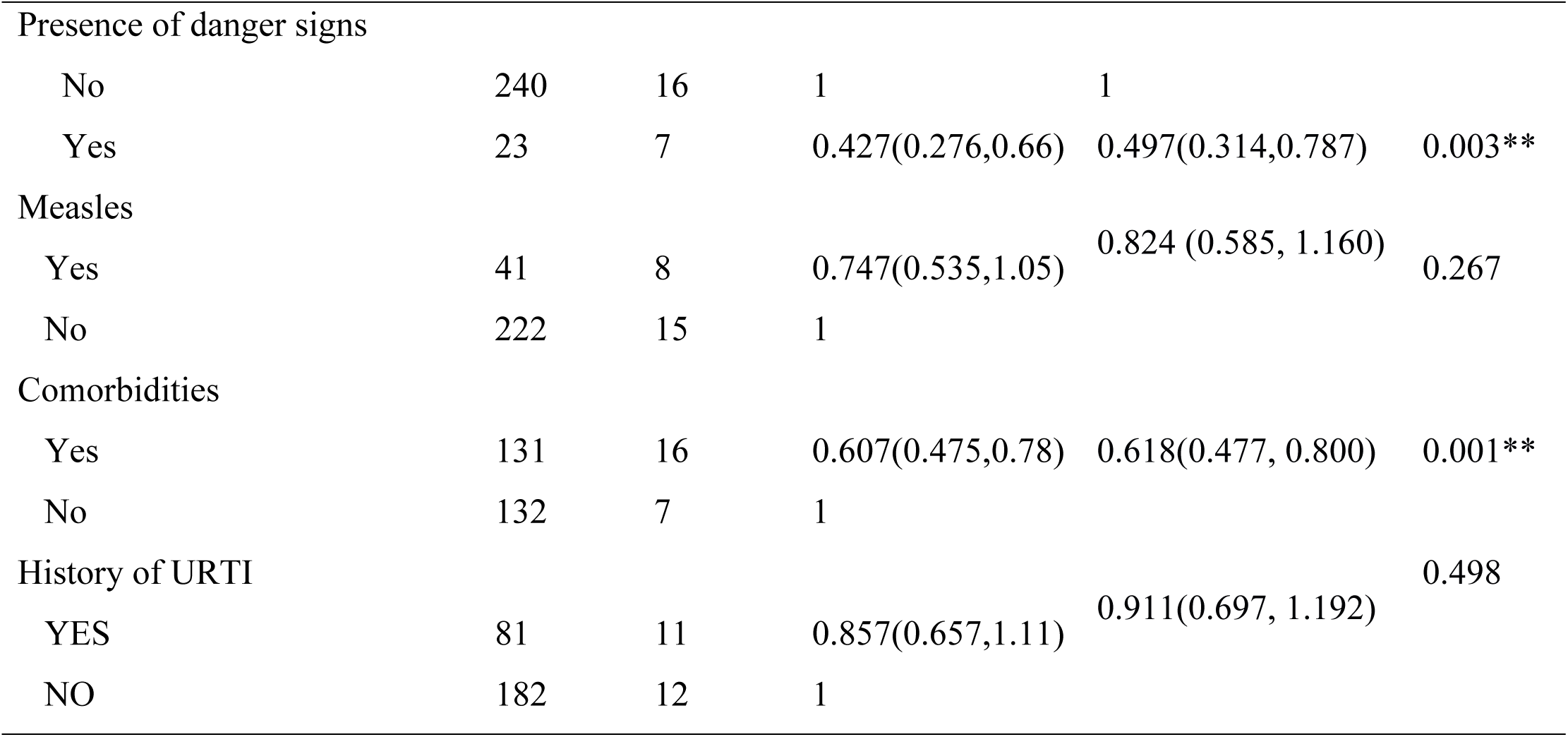
Bivariate and Multivariate cox regression of associated factors of time to recovery of severe pneumonia.

| Variable | Event | Censored | CHR,95% CI | AHR,95% CI | P Value |
| --- | --- | --- | --- | --- | --- |
| Nutritional status |  |  |  |  |  |
| Well nourished | 118 | 2 | 1 | 1 |  |
| Mild acute malnutrition | 87 | 4 | 1.046(0.79,1.38) | 1.030(0.776,1.367) | 0.838 |
| Moderate acute malnutrition | 35 | 6 | 0.59(0.40,0.872) | 0.655(0.443,0.968) | 0.034** |
| Severe acute malnutrition | 23 | 11 | 0.45(0.288,0.711) | 0.567(0.357, 0.90) | 0.016** |
| Immunization status |  |  |  |  |  |
| Fully immunized | 119 | 3 | 1 | 1 |  |
| Partially immunized | 47 | 3 | 0.96(0.68,1.34) | 1.048(0.738, 1.489) | 0.792 |
| Immunized at age | 83 | 10 | 1.04(0.78,1.38) | 1.027(0.773,1.365) | 0.853 |
| Not immunized | 14 | 7 | 0.4(0.23,0.708) | 0.616(0.342, 1.110) | 0.107 |
| Housing condition |  |  |  |  |  |
| Separated from kitchen | 161 | 8 | 1 | 1 |  |
| Not separated from kitchen | 102 | 15 | 0.811(0.632,1.04) | 1.062(0.816,1.381) | 0.656 |
| Breastfeeding status |  |  |  |  |  |
| Exclusively breast feeding | 183 | 11 | 1 | 1 |  |
| Not exclusively breastfeeding | 80 | 12 | 0.84(0.65,1.09) | 0.910(0.696, 1.190) | 0.491 |
| Presence of danger signs |  |  |  |  |  |
| No | 240 | 16 | 1 | 1 |  |
| Yes | 23 | 7 | 0.427(0.276,0.66) | 0.497(0.314,0.787) | 0.003** |
| Measles |  |  |  |  |  |
| Yes | 41 | 8 | 0.747(0.535,1.05) | 0.824 (0.585, 1.160) | 0.267 |
| No | 222 | 15 | 1 |  |  |
| Comorbidities |  |  |  |  |  |
| Yes | 131 | 16 | 0.607(0.475,0.78) | 0.618(0.477, 0.800) | 0.001** |
| No | 132 | 7 | 1 |  |  |
| History of URTI |  |  |  |  |  |
| YES | 81 | 11 | 0.857(0.657,1.11) | 0.911(0.697, 1.192) | 0.498 |
| NO | 182 | 12 | 1 |  |  |

## DISCUSSION

This study showed that the overall median recovery time from SCAP among-under five children admitted with SCAP at a referral and general hospital in the Sidama region was 5 days (IQR = 4- 7 days). This finding was similar to the estimation of the under-five severe pneumonia recovery time that a study conducted at the pediatric ward at Asella Referral and Teaching Hospital was 5 days (IQR= 3-8 days)(25), a governmental hospital in the South West Region, median time to recovery was 5 days (23); public hospitals in Benishangul-Gumuz Region, Ethiopia, the median hospital stay was 5 days (IQR 2–8) (14). Nearly similar with the study conducted at Debre Markos Referral Hospital, median recovery time was 4 days (IQR 3-7)(26); Nigist Eleni Mohammed Memorial Comprehensive Specialized Hospital in Hossana, median time to recovery was 4 days (interquartile range = 3-5) (27), and selected public hospitals in Addis Ababa, the median recovery time was 6 days (24), but higher than that the study conducted at the University of Gondar Comprehensive Specialized Hospital in Ethiopia, median time to recovery was 3 days (IQR= 3–6) (5). This may be due to the study setting, quality of the health care system, and differences of socio-demographic factors.

The incidence of recovery rate was 17.27 (95% CI, 15.3, 19.4) per 100 person-days. This study was nearly similar to the study conducted in Debre Markos referral hospital, North West Ethiopia; the incidence of recovery rate was 16.25 (95% CI: 14.54–18.15) per 100 person-days observation (26), and higher than the study that was conducted at the University of Gondar Comprehensive Specialized Hospital in Ethiopia, where the incidence of recovery rate from severe pneumonia was 13.5 (95% CI: 13.54–17.15) per 100-persons (5), and at a governmental hospital in the South West Region of Ethiopia, with an incidence rate of 12.48 per 100 person-day observations (23). However, the overall incidence of this study was lower than the study conducted at Hossana Nigist Eleni Mohammed Memorial Comprehensive Specialized Hospital, the incidence rate of recovery was 24.16 per 100 person-days (27). This discrepancy could be due to differences in a healthcare setting and in sample size. In our study, the sample size was a smaller sample size than the above studies.

Severe acute malnutrition and moderate acute malnutrition are significantly associated with the recovery time from severe pneumonia. The hazard ratio of recovery of children who were admitted with severe acute malnutrition reduced by 43.3% compared to those admitted with well-nourished. This finding is supported by the study conducted in Gambia (17) and the Benishangul-Gumuz Region, Ethiopia (14). Malnourished children recovered from the disease slower in comparison to normal children. Malnutrition magnifies the effect of disease and causes more severe disease episodes and complications, and people spend more time ill. Under nutrition is often associated with a depressed immune system in children and consequently worsens the prognosis of disease and makes them prone to difficult recovery from pneumonia (28).

The other predictor that has a significant effect on the median time to recovery from SCAP is the presence of danger signs. The hazard ratio of recovery from SCAP was 50.3% lower among patients who had the presence of danger signs (complicated or danger signs) at admission. This finding is in line with other studies conducted at Debre Markos Referral Hospital, North West Ethiopia (29), and Asella Referral and Teaching Hospital, Ethiopia (25). This is because of the severity of the disease. As the disease becomes severe, the required time to recover from the disease becomes too long due to the pathophysiologic state of the disease (30).

The other important predictor that was significantly associated with recovery time from SCAP was the presence of comorbid disease. The hazard ratio of recovery of children admitted with co- morbidity decreased by 38.2% as compared to those children who had no co-morbidity. This finding is in line with the study conducted in Ayder Comprehensive Specialized Hospital, Tigray, Ethiopia (31). This is due to that when children acquire different illnesses at a time or children with comorbidities take a prolonged time of recovery. This may be associated with a decrease in immune system of children and might lead to poor adherence of prescribed drugs due to pill burden, which could cause prolonged hospitalization of children with comorbidities (32,33).

## CONCLUSIONS AND RECOMMENDATIONS

The median recovery time among pediatric patients admitted with severe pneumonia was relatively high. Children who were being malnourished, the presence of danger signs, and the presence of comorbidities were factors that were associated with time to recovery. The hospitals and other stakeholders should facilitate to the parents and community at large regarding the benefit of nutrition, early detection, and treatment of the danger signs and comorbidities by preparing a schedule for a health education session. Harmonized and integrated intervention needs to be taken to reduce prolonged hospitalization from severe pneumonia by nutrition, early detection and treatment of danger signs, and preventing and early detection of comorbidity diseases.

## Data Availability

data avavailable at authors.

## Acronyms and Abbreviations

AHR: adjusted hazard ratio
BSC: bachelor science
CHR: crude hazard ratio
CI: confidence interval
HIV: human immune virus
HUCSH: Hawassa university comprehensive specialized hospital
IQR: interquartile range
NICU: neonate intensive care unit
SAM: severe acute malnutrition
SCAP: severe community acquired pneumonia
URTI: upper respiratory tract infection
WHO: world health organizations

## Ethical approval and informed consent

Ethical clearance for this study was obtained from the Institutional Review Board of the College of Medicine and Health Science, School of Public Health, Hawassa University. A support letter from the school was submitted to HUCSH administration, Adare General Hospital, Tula General Hospital, Leku General Hospital, Yirgalem General Hospital, and Bona and Daye General Hospital Administration to get permission prior to actual data collection. Written informed consent was taken from the Ethics Committee of Referral and General Hospital in the Sidama region prior to study initiation. The information was gathered from face-to-face interviews after taking both written and verbal informed consent from parents or guardians and medical records, which are confidential, secured, and used for research purposes only.

## Consent for publication

Not applicable

## Availability of data and materials

Readers who wish to gain access to the data can write to the corresponding author: Fantahun funte at

## Authors’ Contributions

All authors made a significant contribution to the study work, reported, whether that is in the conception, study design, execution, acquisition of data, analysis, and interpretation, or all these areas; took part in drafting, revising, or critically reviewing the article. They have agreed on the journal to which the article has been submitted and have agreed to be accountable for all aspects of the work.

## Acknowledgements

Firstly, our thanks go to Hawassa University College of Medicine and Health Science, Hawassa University, Office of the Vice President for Research and Technology Transfer Research Programs Directorate. Our sincere appreciation also extends to the Sidama Regional State Health Bureau and Hawassa City Health Bureau for their invaluable collaboration and support during data collection. We would also like to express heartfelt gratitude to the dedicated data collectors for their tireless efforts and contributions to the data collection process, which have been indispensable.

Lastly, heartfelt thanks go out to the study participants for their willingness to provide crucial information that has made this study possible. Their contribution is invaluable, and we were deeply appreciative of their trust and participation.

## Authors’ information

Fantahun Funte Ware (MPH); Nana Chea Hankalo (MPH, Assistant professor, Hawassa university, college of medicine and health science); Ruth Abriham Tadesse (MPH, lecturer at Hawassa university, college of medicine and health science).

## Conceptual frame work

The conceptual framework of severe pneumonia under five years old involves various factors that influence time to recovery. Some of these factors include age, underlying health conditions, residence, nutritional factors, immunization, factors related to duration of illness and existing other medical conditions and/or co-morbidities.

**Figure 1.**
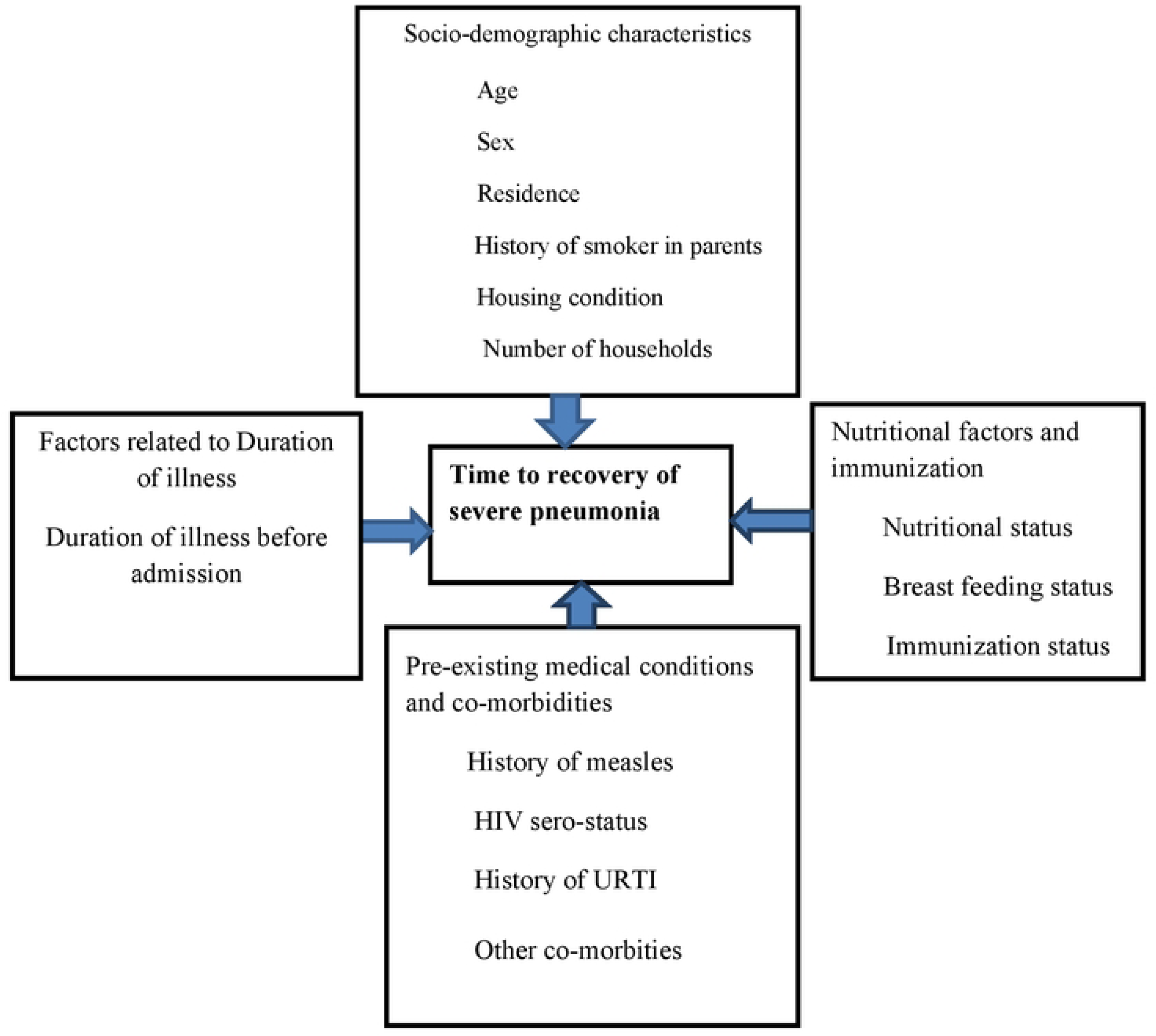
Conceptual framework of time to recovery and associated factors of time to recovery of severe pneumonia under five years adapted from different studies, Conceptual framework to assess risk factors of acute respiratory infection among under-five children (12,25,28,42).

**Figure 2.**
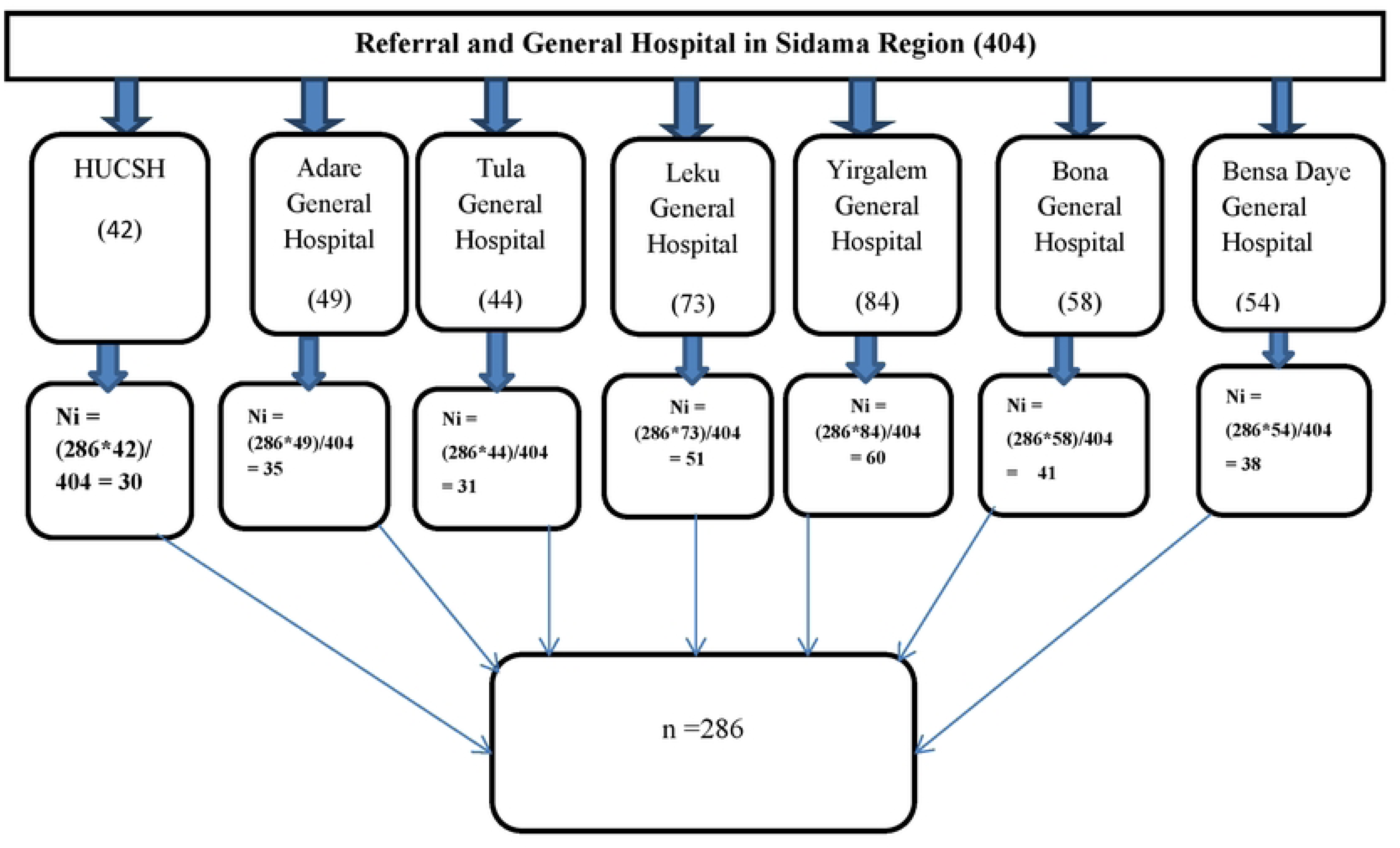
Proportional allocation of sample size for each hospital in Sidama Region.

## Treatment outcome of severe pneumonia

An1ong 286 cases analyzed of which 11(3.8%) were died; 263(92%) were irnproved and 12(4.2%) had unknown outcomes (self against medical services and referred).

**Figure 3.**
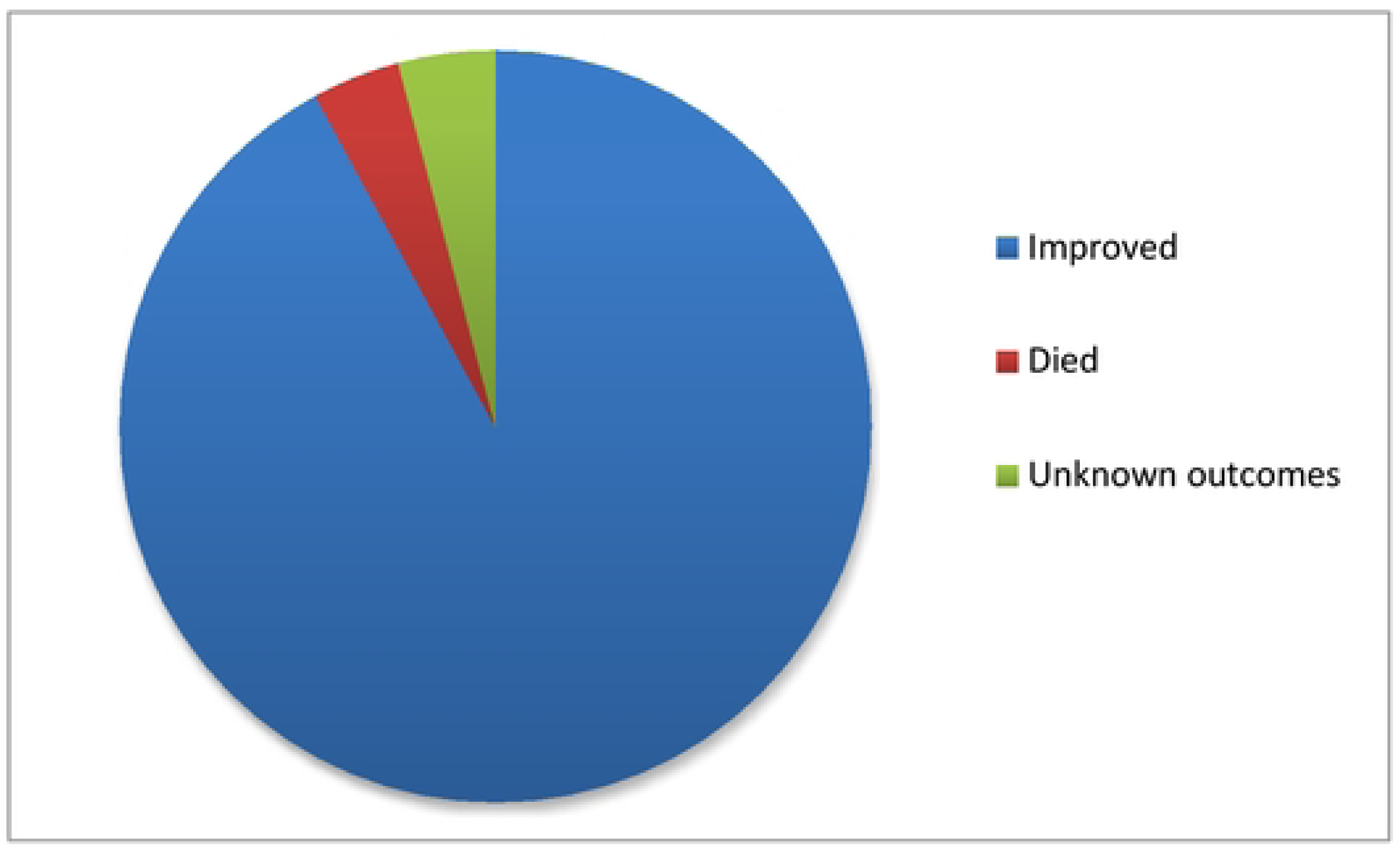
Treatment outcome of severe pneumonia of under five children in referral and general hospitals in Sidama Region (N=286).

## Kaplan-Meier Survival Curve and survival estimate

The median survival time to recovery from severe pneumonia among under five children was found to be 5 days (lQR: 4-7). The incidence of recovery rate was 17.27 (95% CI, 15.3, 19.4) per 100 person days. The total person days contributed by the study participants were 1523 days. The chance of survival was higher on the first day after a severe pneumonia diagnosis, but it decreased as follow up time increased.

**Figure 4.**
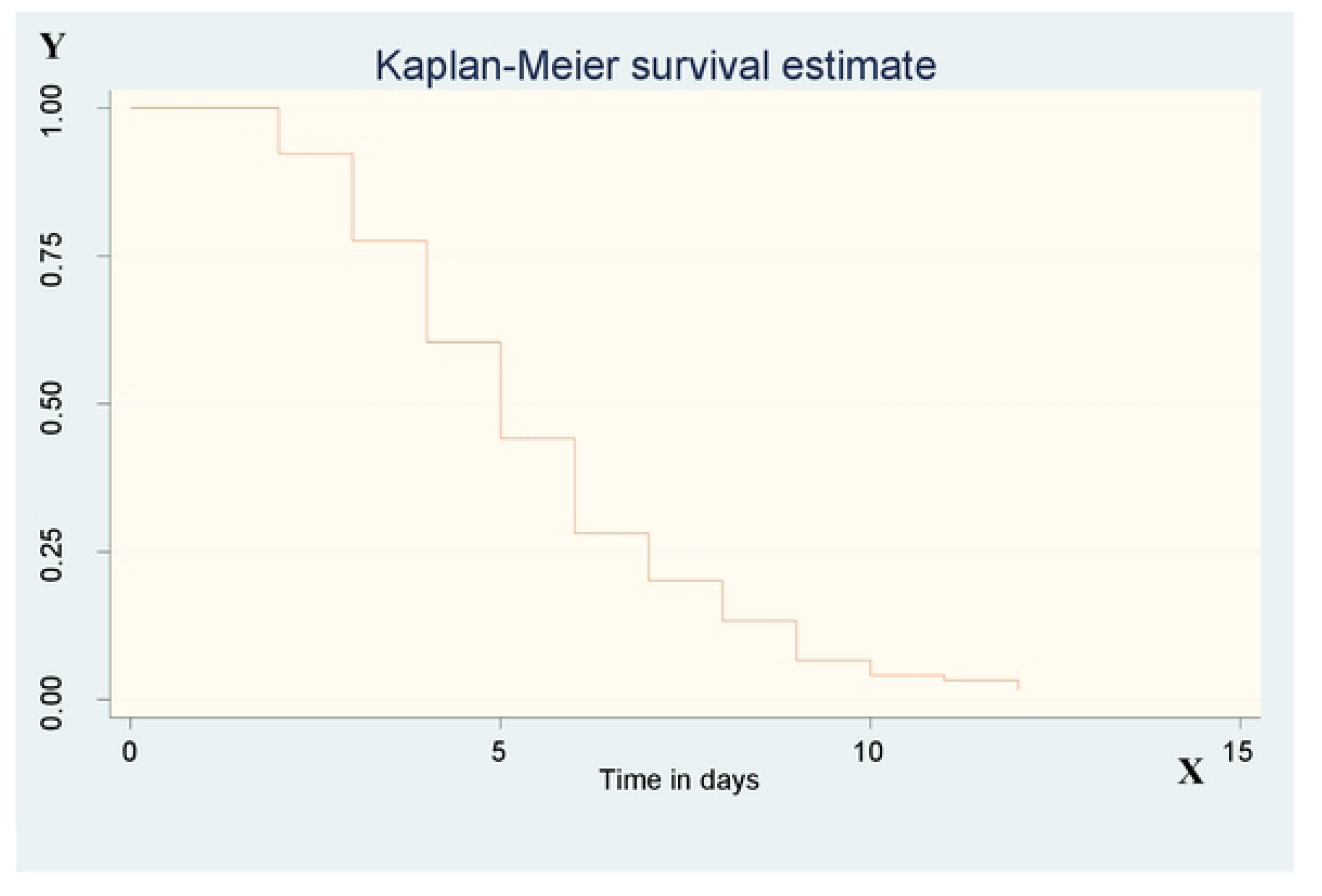
Overall Kaplan-Meier estimation of survival function from admission to recovery among under five severe pneumonia children admitted at the referral and general hospital in Sidama region, Ethiopia, 2024.

## Survival function and comparison of survival functions for different categorical Variables

The Kaplan-Meier estimator survival curve provides the survivor function estimate across several sets of covariates to facilitate comparisons. As was shown below, distinct graphs containing the estimates of the Kaplan-Meier survivor functions were created for various categorical covariates. When two survival functions lie above one other, it often indicates that the group described by the upper curve has a higher survival rate or has had a more favorable experience with survival than the group indicated by the lower curve.

**Figure 5.**
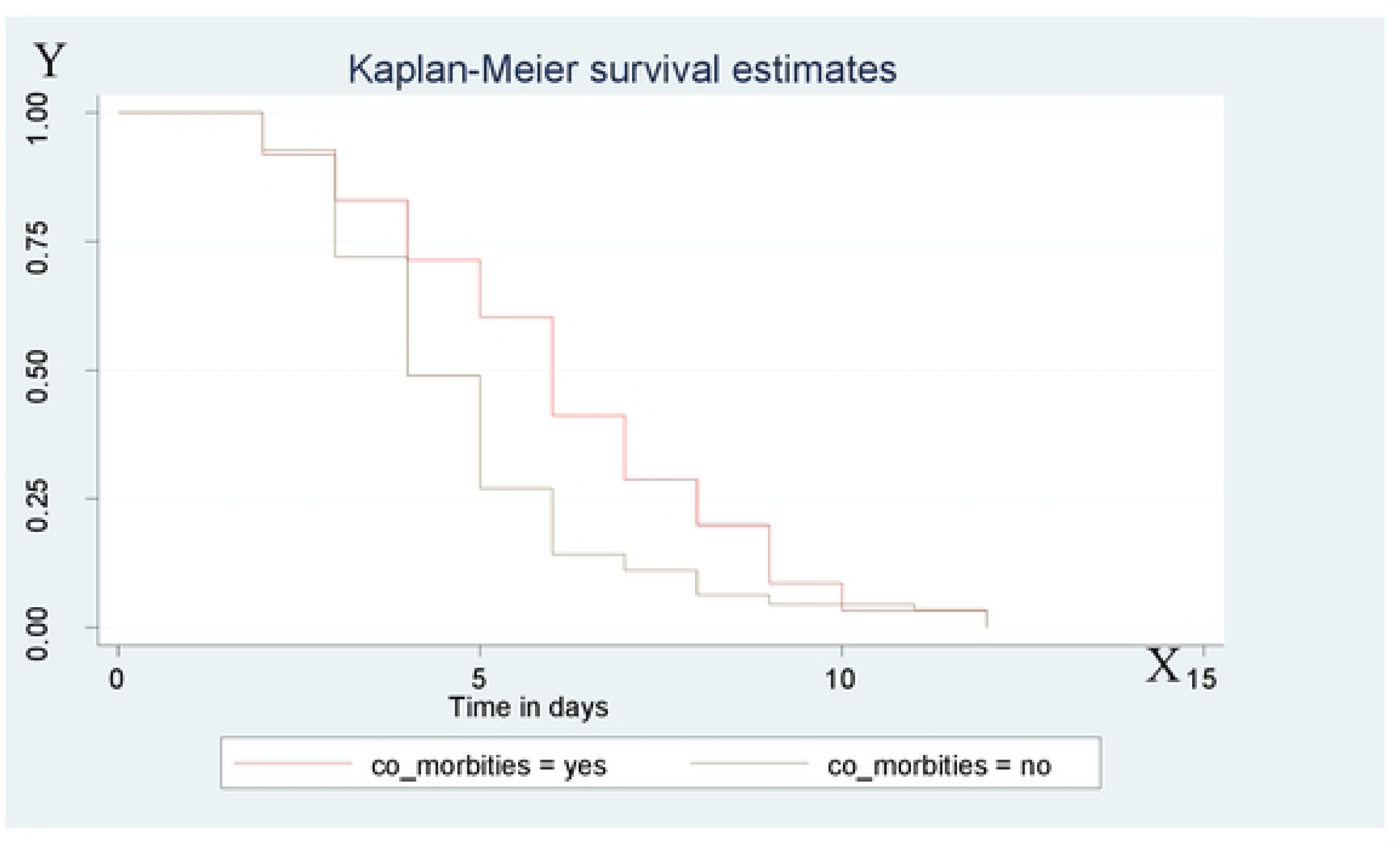
Kaplan-Meir survival estimate among under five children admitted by severe pneumonia at referral and general hospitals in Sidama region, Ethiopia, with the category of comorbiditiess.

**Figure 6.**
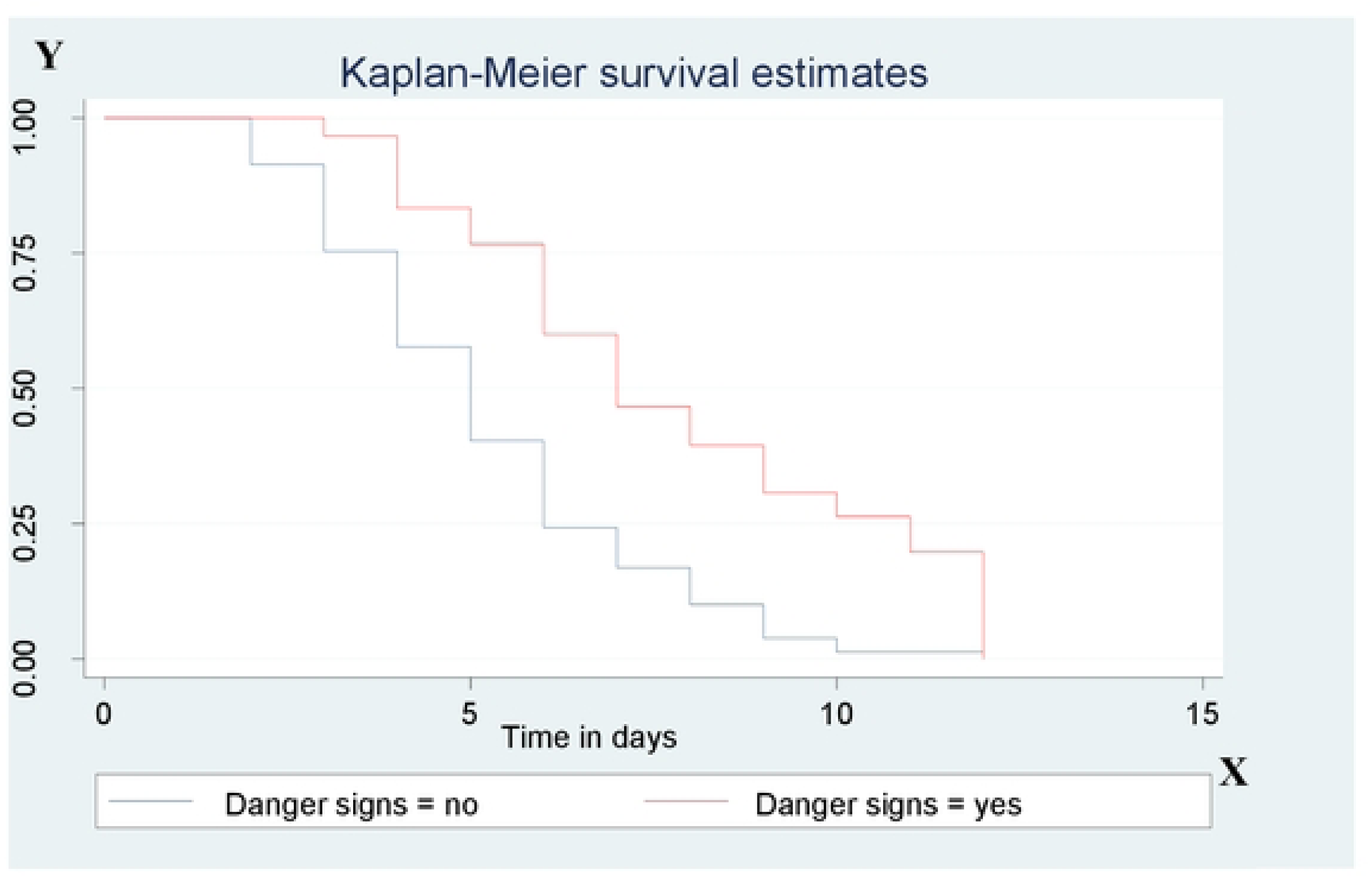
Kaplan-Meir survival estimate among under five children admitted by severe pneumonia at referral and general hospitals in Sidama region, Ethiopia, with the category of presence of danger signs.

**Figure 7.**
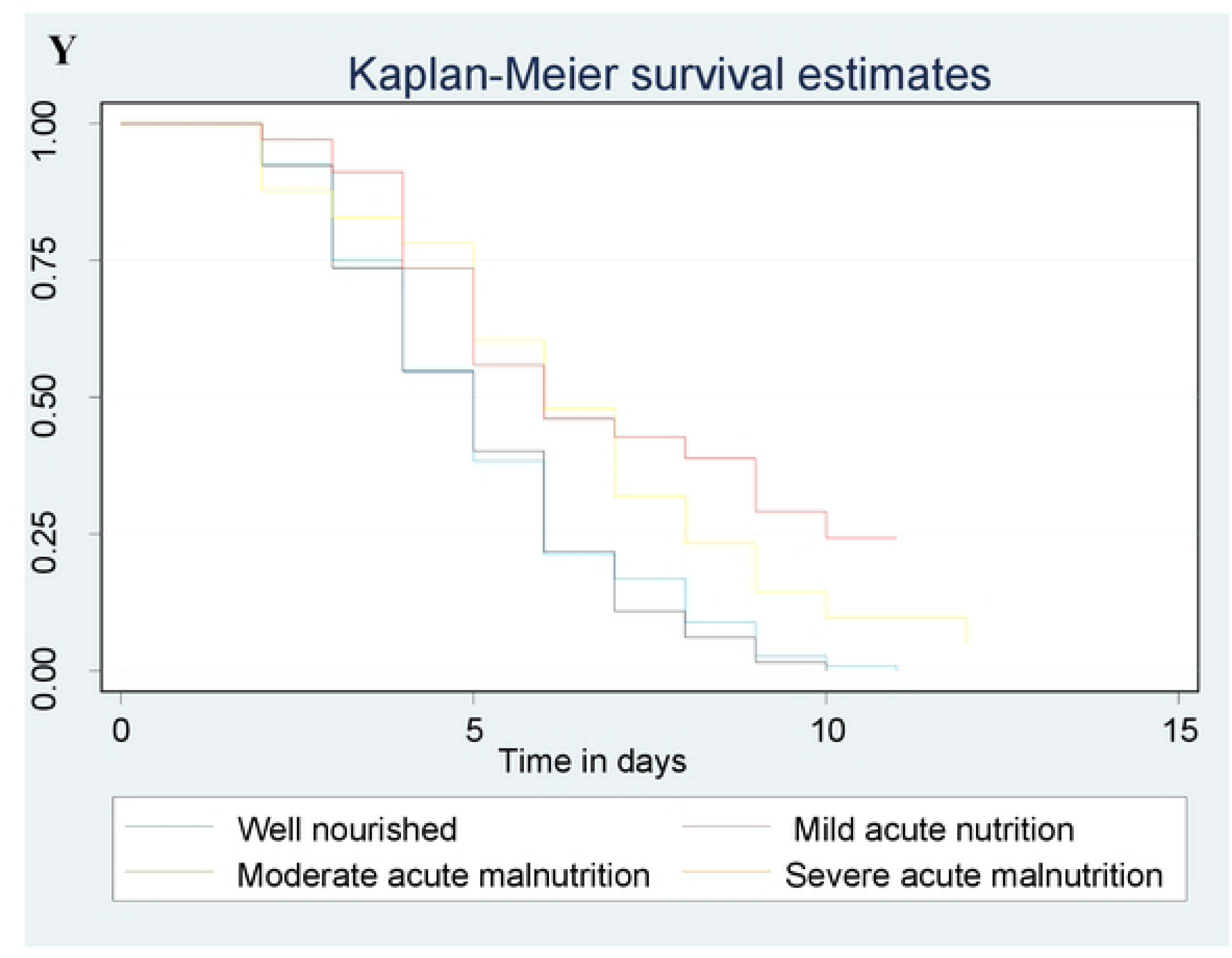
Kaplan-Meir survival estimate among under five children admitted by severe pneumonia at referral and general hospitals in Sidama region, Ethiopia, with the category of Nutritional status.

